# Post-acute COVID-19 syndrome and its prolonged effects: An updated systematic review

**DOI:** 10.1101/2021.05.09.21256911

**Authors:** Jahanzeb Malik, Syed Muhammad Jawad Zaidi, Raafe Iqbal, Kashif Khan, Muhammad Ali, Abdul Sattar Rana, Ali Umer Waqar, Uzma Ishaq

## Abstract

**Objective:** This systematic review aimed at estimating the demographics, clinical characteristics, and prevalence of post-acute COVID-19 symptoms in view of published literature that studied prolonged clinical manifestations after recovery from acute COVID-19 infection.

**Methods:** After protocol setting, relevant articles were searched on various databases including PubMed, Medline, the Cumulative Index to Nursing and Allied Health Literature (CINAHL), Embase, and Web of Sciences using MeSH keywords.

**Results:** Out of the 153 articles reviewed, 21 articles qualified for the final analysis. The most common persistent clinical manifestations were fatigue (54.11%), dyspnea (24.38%), alopecia (23.21%), hyperhidrosis (23.6%), insomnia (25.98%), anxiety (17.29%), and arthralgia (16.35%). In addition to these symptoms, new-onset hypertension, diabetes, neuropsychiatric disorders, and bladder incontinence were also reported.

**Conclusion:** Clinical features of post-acute COVID-19 infection can manifest even after 60 days of initial infection. Multidisciplinary care along with regular follow-up must be provided to such patients.

## Introduction

Severe acute respiratory syndrome coronavirus 2 (SARS-COV-2) presented as clustered cases of atypical pneumonia in the city of Wuhan in the Hubei province of China. In March 2020, coronavirus disease 2019 (COVID-19) was declared as a pandemic by World Health Organization (WHO) and since then approximately 148 million people have been infected with the virus [1]. There is a well-established pool of scientific knowledge about the acute effects of COVID-19 and unprecedented efforts of the scientific community have now shifted towards the long-lasting sequelae of the disease, the real-world effects of which are yet to be seen [2-5].

Prolonged symptoms and signs are being reported in observational studies and case reports every day. Although such symptoms are usually experienced in survivors of critical illness, the post-acute effects of COVID-19 are equally being reported in patients with mild severity of disease who do not require hospitalization [6,7]. No established randomized controlled trials have assessed predictors of post-acute COVID-19 and many observational studies are underpowered with a small sample size to produce a significant effect model and a pooled prevalence on a global scale. Therefore, this systematic review was conducted for published research articles to estimate the incidence of abnormal manifestations after recovery from the acute COVID-19.

## Methods

### Protocol development and selection criteria

A protocol for the selection of articles and carrying out the systematic review of the literature was made after a consensus among the authors and subject experts but it was not deposited in a registry. Data was collected after protocol approval from the ethical review board of Foundation University Medical College (ID#FFH/ADC/021/21).

### Search strategy and data extraction (selection and coding)

The main databases used for study selection were PubMed and Medline through LitCOVID [8], the Cumulative Index to Nursing and Allied Health Literature (CINAHL), Embase, and Web of Science. Articles published before 1st May 2021 were included in the search. We included randomized clinical trials, observational, cross-sectional, and cohort studies which were in the English language, and peer-reviewed published articles that reported signs and symptoms after at least two weeks from the recovery of acute COVID-19. Only studies with more than 50 participants were included. Post-acute COVID-19 syndrome was defined as symptomatology after two weeks of recovery from COVID-19. Pre-prints, case reports, editorials, and data notes were excluded. After the initial search and removal of duplicates, all the search was imported on EndNote version 20 (Clarivate Analytics™). All the screening and inclusion of the articles were conducted by two independent reviewers (ASR, SMJZ) blinded to each other’s decisions. Once the initial screening was finished, all the included studies were referenced in Mendeley. The two reviewers (ASR, SMJZ) reviewed full texts for final inclusion. Where there was a dispute, a third reviewer (MA) resolved it between them. The descriptive variables extracted were country, setting, follow-up time, sample size, mean age and percentage of gender, outcomes, symptoms, and signs, and names used for post-acute COVID-19 syndrome.

The search terms used in the search strategy were as follows: (((“long-haul” coronavirus disease OR post-acute COVID-19 OR “convalescent” COVID-19) OR prolonged coronavirus infection OR coronavirus disease [Mesh]) OR “severe acute respiratory syndrome coronavirus 2” chronic disease [Supplementary Concept]) OR recurrent OR lingering OR complications of “COVID-19” [Mesh] OR “betacoronavirus” [Mesh])) AND 2019/12 [PDAT]: 2030 [PDAT]))). The systematic review followed the Preferred Reporting Items for Systematic Reviewers and Meta-analysis (PRISMA) guidelines [9] and the PRISMA flowchart is demonstrated in **Figure 1**.

**Figure 1.**
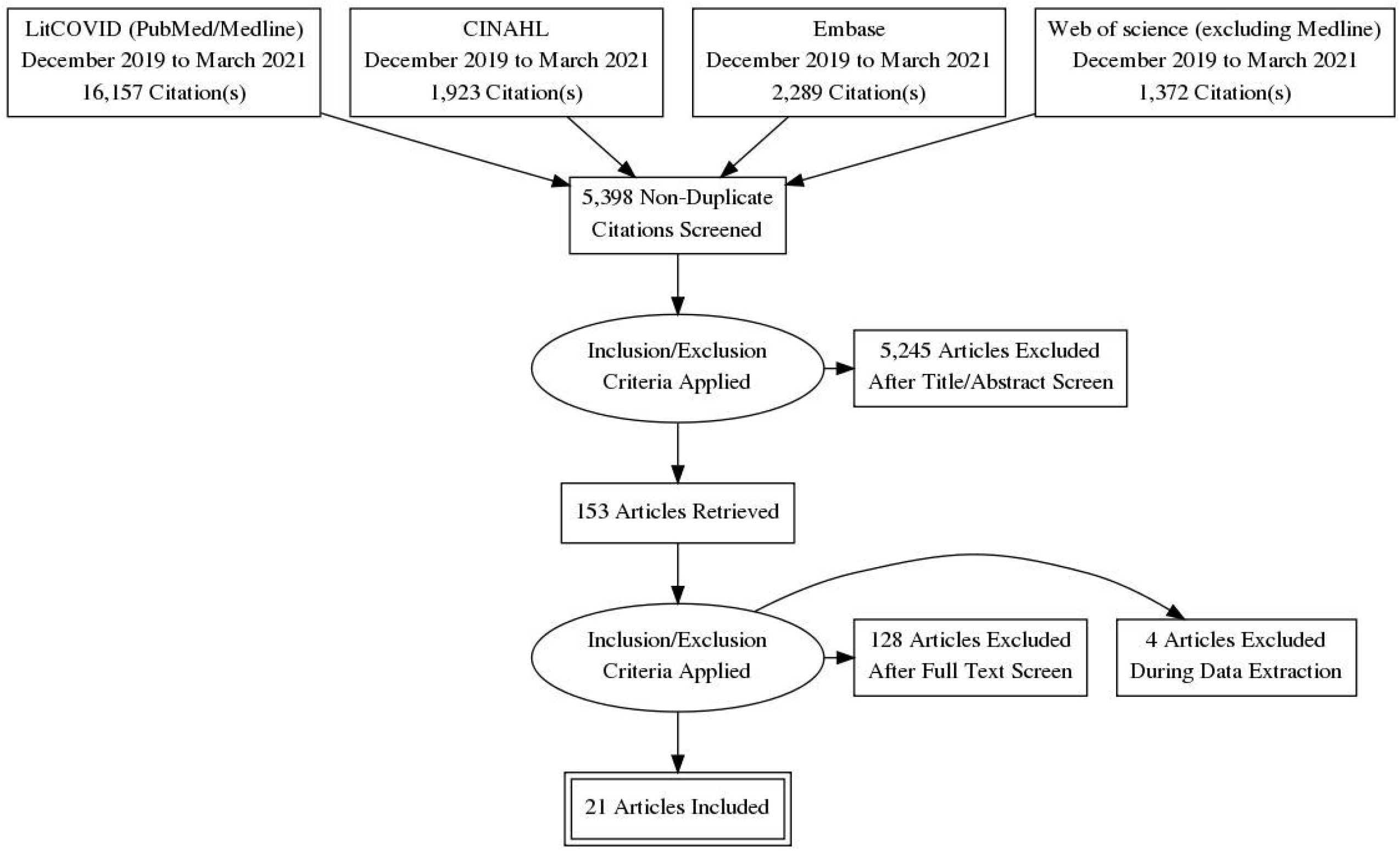
PRISMA flow chart.

### Risk of bias assessment

All included articles were assessed using the National Institute of Health quality assessment tool for clinical studies and Newcastle-Ottawa Scale (NOS) [10]. Scales are provided as **supplementary resources 1 and 2. Table 1** enumerates the quality status of the included studies.

**Table 1.** Quality assessment of included articles according to NIH quality assessment tool and Newcastle-Ottawa scale for observational studies.

| Judgement | Clinical studies |  |
| --- | --- | --- |
|  | NIH quality assessment tool; n (%) | Newcastle-Ottawa scale; n (%) |
| Good | 8 (38%) | 10 (47.61%) |
| Fair | 12 (57.14%) | 11 (52.38%) |
| Poor | 1 (4.76%) | 0 |

### Strategy for data synthesis

For statistical analysis, Statistical Package for the Social Sciences (SPSS) version 26 (IBM Corp. Armonk, NY, USA) was used, and based on the distribution of quantitative variables, they were expressed as mean ± standard deviation (SD) or median and interquartile range (IQR). Qualitative variables were presented as frequency (n) and percentages (%).

## Results

A total of 21,741 titles and abstracts were screened for this review. Of these, 153 full texts were reviewed and according to the review protocol, 56 were excluded because of inappropriate sample size, 47 presented acute COVID-19 symptoms, 23 were case series, and 6 excluded as data notes. A total of 21 studies were included for final analysis and review and their general characteristics are shown in **Table 2**. Many studies assessed a specific long-term symptom after COVID-19 recovery and the PRISMA flowchart for study selection is presented in **Figure 1**. A total of 10 studies were from Europe and three from the United States. Others were from Mexico, Saudi Arabia, China, Australia, and Bangladesh. All studies included were on either previously hospitalized or non-hospitalized patients, and most of them had mild, moderate, and severe states of COVID-19 patients. Total follow-up time was more than one month in the majority of the studies and the number of the patient cohort was 54,730 participants with a median age of 54 years. Except for two studies, there was no stratification among gender differences between post-COVID-19 symptoms.

**Table 2.** General article characteristics and study population. Standard deviation (SD), interquartile range (IQR)

| # | Author [ref] | Country | Setting | Follow-up<br>(Mean) | Study participants | Sample size<br>(n) | Age; mean ±<br>SD/median<br>(IQR) | Males; % | Outcome variables |
| --- | --- | --- | --- | --- | --- | --- | --- | --- | --- |
| 1 | Carfi et al.[14] | Italy | Single-centered | 60.3 ± 13.6<br>days after onset | Patients meeting the<br>following criteria (no | 143 | 56.5 ± 14.6 | 93 out of 143<br>(62.9%) | Quality of life assessment<br>after acute COVID-19, |
|  |  |  |  | of symptoms | fever for 3 consecutive days, improvement in symptoms, and 2 negative test results for SARS-CoV-2 virus 24 hours apart) |  |  |  | length of hospital stay, Number of persistent symptoms. Fever, fatigue, red eyes, chest pain, cough, anosmia, dysgeusia, myalgia, diarrhea |
| 2 | Mandal et al. [16] | London, United Kingdom | Multi-centric (3 hospitals) | 4-6 week after discharge | Patients with abnormal blood tests or imaging at discharge. | 384 | 59.9 ± 16.1 | 62% | Symptom persistence including breathlessness, cough, fatigue, and, poor sleep quality. Laboratory parameters including TLC, platlet count, Lymphocyte count, D dimers, LFTs, and CRP levels |
| 3 | Chopra et al.[21] | United States | Multi-centric (38 hospitals) | 60 days post discharge | ICU/Hospitalized COVID-19 patients discharged between 16 March and 1 July 2020 at 38 hospitals. | 488 | 62 (50-72) years | 51.8% | Mortality and rehospitalization, Primary care follow-up, New/worsened symptoms, Return to normal activity, Emotional impact, Financial loss/impact |
| 4 | El Sayed et al.[17] | Saudi Arabia | Single centered | After discharge. Duration not mentioned | Patients of COVID-19 after 2 consecutive negative PCR tests attending pulmonology clinic for<br>follow-up | 200 | 36.58 ± 9.85 | 114 out of 200 (57%) | Assessment of fatigue and anhedonia using validated scales. |
| 5 | Mahmud et al.<br>[6] | Dhaka,<br>Bangladesh | Single centered | One month<br>after discharge | Discharged COVID-19<br>patients | 355 | 39.8 ± 13.4 | 207 out of<br>355 (58.3%) | The frequency and interval<br>of a spectrum of post<br>COVID-19 symptoms were<br>assessed. These include<br>post viral fatigue, persistent<br>cough, insomnia, Circadian<br>rhythm sleep disorders,<br>headache, vertigo, Post-<br>exertional dyspnea, rash,<br>pneumonia, restless leg<br>syndrome, chest pain,<br>Adjustment disorder, Nasal<br>blockage, Excessive<br>sweating, Disturbance of<br>memory, New-onset<br>diabetes or hypertension,<br>myalgias, and Precipitation<br>of gout |
| 6 | Carvalho-Schneider et al.<br>[28] | France | Single centered | Two months<br>after onset of<br>symptoms. | Post COVID-19 patients<br>with or without clinical<br>signs of pneumonia but<br>without a<br>need for oxygen therapy (mild/moderate disease) | 150 | 49 ± 15 years | 44% | Persisting symptoms at Day<br>30 and 60 which included<br>Fever, dyspnea, chest pain,<br>abnormal auscultation, flu-<br>like symptoms, digestive disorders, weight loss, anosmia, palpitations, arthralgia, cutaneous rashes |
| 7 | Marwa et al. [36] | Egypt | Single centered | Duration not mentioned | Patients recovered from COVID-19 | 287 | 32.3 ± 8.5 | 103 out of 287 (35.8%) | Fatigue, anxiety, joint pain, continuous headache, chest pain, dementia, depression, dyspnea, blurred vision, tinnitus, intermittent fever, obsessive compulsive disorder |
| 8 | Galván-Tejada et al. [32] | Mexico | Multi centeric | Duration not mentioned | Cases: Patients who had a laboratory-confirmed diagnosis of SARS-CoV-2, and in whom at least fourteen days have passed since the appearance of symptoms.<br>Controls: Patients with no laboratory or clinically proven COVID-19 infection | 141 cases and 78 controls. (Total 218) | Means of 39.14 years for females and 39.01 for males respectively | 49% | Fever, myalgia, rhinorrhea or coryza, asthenia, cough, cephalgia, red eyes, odynophagia, nausea, vomit or diarrhea, anosmia or dysgeusia, stomach pain or discomfort, dyspnea, chills |
| 9 | Moreno-Pérez et al.[18] | Spain | Single centric | 10-14 weeks after discharge | Hospitalized Patients who had laboratory proven SARS-COV-2 | 277 | 56.0 (42.0–67.5) | 52.7% | Post- COVID syndrome. These include pneumonia, fatigue, anosmia, dyspnea, persistent cough, headache fever, diarrhea, neurological symptoms, and laboratory features |
| 10 | Halpin et al. [22] | United Kingdom | Single centered | 4 to 8 weeks after discharge | Hospitalized Patients who had laboratory proven SARS-COV-2 and were discharged from hospital | 100 | For ward patients: 70.5 (20–93) For ICU patients: 58.5 (34–84) | 54% | Fatigue, Breathlessness, Neuropsychological symptoms, Speech and swallowing problems, weight loss/gain, bowel/bladder incontinence, Perceived health, quality of life, and Vocation change since COVID-19 illness. |
| 11 | Huang et al [7] | China | Single centered | Median follow up days: 186 (175-199) days | patients with laboratory confirmed COVID-19 who were discharged between Jan 7, and May 29, 2020 | 1733 | 57.0 (47.0–65.0) | 52% | Fatigues, sleeping problems, hairloss, anosmia, palpitations, joint pain, decreased appetite, taste disorder, chest pain, myalgias, rashes, swallowing difficulty, Low grade fever, eGFR, and quality of life |
| 12 | Xiong et al. [27] | China | Single centered | Follow up time of more than 3 months for | All COVID-19 survivors who were diagnosed with COVID-19 | 538 | 52.0 (41.0-62.0) years | 245 out of 538 (45.5%) | Fatigue, swelling, myalgias, arthralgia, chills, limb edema, dizziness, chest |
|  |  |  |  | each survivor | according to WHO interim guidance and were discharged from the hospital by 1 March 2020 |  |  |  | pain, post activity polypnea, cough sputum, throat pain, palpitations, discontinuous flushing, new onset hypertension, depression, anxiety, and alopecia |
| 13 | Tenforde et al. [35] | United States | Single centered | 14–21 days after the test date | adults aged $\geq 18$ years who had a first positive RT-PCR test for SARS-CoV-2, and reported persistence COVID-19 symptoms | 270 | 26% patients aged between 18–34 years, 32% aged between 35–49 years, and 47% aged $\geq 50$ years | 130 out of 270 (48.14%) | Risk Factors for Delayed Return to Usual Health<br>Among COVID-19 patients were evaluated. The outcome variables included age, comorbidities, ethnicity, gender |
| 14 | Taquet et al. [24] | United States | Multicentric, electronic records | 14-90 days | Discharged COVID-19 patients with no previous psychiatric illness | 44,779 | 49.3 (19.2) | 45.1 | New onset psychiatric illness<br>disorders psychotic, insomnia, mood disorders (depressive episodes) anxiety disorders (PTSD, panic disorder, adjustment disorder and generalized anxiety disorder). |
| 15 | Townsend et al.[15] | Ireland | Single centered, outpatient clinic | 56 days to 12 weeks | Mild, moderate Symptomatic patients and Hospitalized patients | 128 | 49.5 ± 15 years | 46.1% | Persistent fatigue |
| 16 | Garrigues et al. [37] | France | Single centered | 110.9 days | Discharged COVID-19 patients who were Hospitalized in ward or ICU | 120 | 63.2 (15.7) years | 75 out of 120 (62.5%) | Cough, chest pain, fatigue, dyspnea, ageusia, anosmia, hair loss, attention disorder, memory loss, sleep disorder |
| 17 | Horvath et al. [38] | Australia | Multicentric, computed records | 83 days | Discharged COVID-19 patients with mild to moderate disease intensity | 102 | 45 (17-87) years | 40% | Smell reduction, taste change, cough, fever, headaches, worsening nasal blockage, runny nose, fatigue, sore throat. |
| 18 | Arnold et al.[39] | United Kingdom | Single centered | 28 days | patients ($\geq 18$ years of age) admitted with COVID-1 | 110 | 60 (46–73) years | 56% | Fever, cough arthralgia, myalgias, chest pain, anosmia, diarrhea, abdominal pain, headache, insomnia, deranged blood tests, spirometry and chest C ray |
| 19 | Osikomaiya et al. [40] | Nigeria | Multi- centered | Two weeks after discharge | Discharged COVID-19 patients who were Hospitalized in ward or ICU | 274 | 41.8 ± 11.8 years | 181 out of 274 (66.1%) | Fever, fatigue, weight loss, malaise, cough, dyspnea, chest pain, anosmia, loss of appetite, dizziness, palpitations, insomnia vertigo, dysgeusia |
| 20 | Leth et al. [41] | Denmark | Single centered | 12-week | Hospitalized COVID-19 that were discharged after negative PCR | 71 patients | 58 (48–73) | 43% | Difficulty in concentration, paresthesia's, headache, anosmia, taste impairment, cough dyspnea, expectoration, sore throat, |
| 21 | Sudre et al.[23] | United Kingdom | Multi-center | 3 months | Mobile health app users with PCR positive COVID-19 patients/negative matched controls | 4,182 | 44 (28, 56) | 28.5% | Number of symptoms, duration of symptoms, quality of life, |

The general quality of the studies was assessed using the NIH quality assessment tool and Newcastle-Ottawa scale for observational studies and except one, all studies were graded in at least fair quality articles (**Table 1**). There were 35 post-acute symptoms and signs presented in the study cohort of these articles. They are stratified in **Table 3**. The most common manifestations were fatigue (54.11%), dyspnea (24.38%), alopecia (23.21%), hyperhidrosis (23.6%), insomnia (25.98%), anxiety (17.29%), and arthralgia (16.35%). Thirteen studies reported fatigue and anosmia, 15 dyspnea, 12 chest pain, and 11 non-productive coughs, and 5 studies showed more than one symptom. Apart from constitutional symptoms of COVID-19, personality and sleep disorders, bladder and bowel incontinence, new-onset hypertension, and diabetes were also seen in the included studies.

**Table 3.** Post-acute COVID-19 signs and symptoms after recovery.

| **Clinical characteristics of post-acute COVID-19** | **Studies (n)** | **Number of patients with symptoms (n)** | **Total number of patients (n)** | **Prevalence; %** |
| --- | --- | --- | --- | --- |
| **Fatigue** | 13 | 2412 | 4457 | 54.11% |
| **Hyperhidrosis** | 1 | 127 | 538 | 23.6% |
| **Migraine-like Headache** | 8 | 221 | 3006 | 0.03% |
| **Vertigo** | 2 | 11 | 629 | 1.74% |
| **Alopecia/Telogen effluvium** | 3 | 537 | 2313 | 23.21% |
| **Dyspnea** | 15 | 790 | 3242 | 24.38% |
| **Anosmia** | 13 | 497 | 3924 | 12.66% |
| **Dry eyes** | 1 | 21 | 146 | 14.38% |
| **Blurred vision** | 3 | 70 | 838 | 8.35% |
| **Dysgeusia/Ageusia** | 7 | 293 | 3009 | 9.73% |
| **Arthralgia** | 5 | 198 | 1211 | 16.35% |
| **Myalgias** | 9 | 204 | 3527 | 5.78% |
| **Adjustment disorder** | 1 | 2 | 355 | 0.56% |
| **Anxiety** | 3 | 160 | 925 | 17.29% |
| **Dementia** | 1 | 82 | 287 | 28.57% |
| **Dizziness** | 3 | 123 | 2467 | 4.98% |
| **Depression** | 2 | 117 | 825 | 14.18% |
| **Cough** | 11 | 434 | 2527 | 17.17% |
| **Expectoration** | 1 | 16 | 538 | 2.97% |
| **Insomnia** | 6 | 725 | 2790 | 25.98% |
| **Obsessive compulsive disorder** | 1 | 14 | 287 | 4.87% |
| **New-onset hypertension** | 3 | 10 | 1180 | 0.84% |
| **New-onset diabetes** | 2 | 3 | 642 | 0.46% |
| **Palpitations** | 5 | 252 | 2952 | 8.53% |
| **Discontinuous flushing** | 1 | 26 | 538 | 4.83% |
| **Restless leg syndrome** | 1 | 2 | 355 | 0.56% |
| **Pedal edema** | 1 | 14 | 538 | 2.6% |
| **Memory disturbances** | 4 | 69 | 849 | 8.12% |
| **Rash/Cutaneous signs** | 4 | 87 | 2417 | 3.59% |
| **Chest pain** | 12 | 459 | 4422 | 10.37% |
| **Sore throat/odynophagia** | 6 | 150 | 2886 | 5.19% |
| **Bowel/bladder incontinence** | 1 | 8 | 100 | 8% |
| **Tinnitus** | 1 | 48 | 287 | 16.72% |
| **Weight loss** | 3 | 42 | 504 | 8.33% |
| **Diarrhea/Vomiting/gastrointestinal issues** | 7 | 236 | 2911 | 8.1% |
| **More than one symptom** | 5 | 456 | 1533 | 29.74% |

## Discussion

The most important pathophysiological mechanism in acute COVID-19 is direct viral toxicity leading to endothelial damage and microvascular injury. This can cause immune system dysregulation and hyperinflammatory states, hypercoagulability, and downregulation of the angiotensin-converting enzyme 2 (ACE2) pathway [11]. In contrast to this, the post-acute effects of COVID-19 are an overlap of phylogenetic similarities with SARS-COV-1 and Middle-Eastern respiratory syndrome (MERS) viruses [12]. However, SARS-COV-2 has a higher affinity for ACE2 compared with SARS-COV-1, and this mechanism may be the contributing factor in the widespread transmission of SARS-COV-2. Furthermore, potential mechanisms behind post-acute symptoms and signs in COVID-19 recovered patients seem multifactorial: the pathophysiologic changes caused by the virus, inflammatory and immune-mediated cell damage, and sequelae of recovery from critical illness [13].

This systematic review demonstrated that 68% of patients have at least one post-acute symptom after recovery from COVID-19. A total of 21 studies were included in this review which fulfilled our inclusion criteria and overall 35 signs and symptoms of post-acute COVID-19 were identified in this cohort of patients (Table 3). The most common symptoms were fatigue, dyspnea, hyperhidrosis, dementia, depression, alopecia, and cough. The majority of presenting symptoms or signs were similar to the acute presentation of COVID-19. However, a possibility remains for other effects to be identified later on in this pandemic. In the following discussion, we will elaborate on the most common symptoms and signs of post-acute COVID-19 to understand each disease in more detail.

Overall, the most common symptom among all the included patients was the feeling of tiredness or fatigue (54.11%) [6,7,14-18]. It was present after three months’ follow-up in critical COVID-19 patients admitted to intensive care units (ICUs) [15]. This phenomenon has been established in survivors of critical illness (post-ICU syndrome), even after years of recovery, where half the patients report symptoms of chronic fatigue syndrome, including incapacitating fatigue, generalized body pain, neurocognitive disturbances, insomnia, and increased sympathetic drive [19]. Viruses like Epstein-Barr virus, cytomegalovirus, and herpes virus have been implicated in causing chronic fatigue syndrome and this review adds SARS-COV-2 as the causative agent of chronic fatigue [20].

Neuropsychiatric symptoms are also reported in some studies, including headache, insomnia, anxiety, depression, bladder and bowel incontinence, ageusia, migraine, and dementia [6,16,21-24]. Similar to chronic fatigue syndrome, the etiology, and pathophysiology of neuropsychiatric symptoms in COVID-19 are multifactorial and unclear. In a cohort of 355 patients in Bangladesh, and 143 patients in Italy, a cumulative 63% of the patients were screened positive in at least one of the domains evaluated for neuropsychiatric sequelae (depression, anxiety, insomnia, obsessive-compulsive disorders, etc.) [6,14]. Clinical depression and anxiety were reported in approximately 17% of patients following COVID-19 [6]. Memory loss in the form of dementia and ageusia is also reported in a few studies, including cognitive impairment with or without fluctuations [25]. All these symptoms could be related to the social stigma of contracting a potentially fatal illness, some effects of sedatives in critical COVID-19 patients with delirium, and hypercoagulability leading to cerebrovascular disease. In addition, post-recovery sleep disturbances can also precipitate psychiatric disorders [26]. Mental health assessment and mental health attention models are very important in the post-acute COVID-19 stage, as they can contribute to a better quality of life in this cohort. Telogen effluvium and alopecia are also reported in three studies, which is defined as temporary hair loss due to excessive shedding of Telogen hair after COVID-19. Although self-limiting, this condition can cause emotional distress in many patients [27].

Dyspnea (24.38%) and cough (17.17%) were the most prominent pulmonary symptoms in this review [28]. Several studies have demonstrated persistent high resolution computed tomography (HRCT) lung abnormalities after 60 days from the initial presentation [29]. In addition, previous studies have exhibited lung dysfunction in more than 50% of the patients compared to our study cohort [7,30]. A decreased diffusion capacity due to loss of lung volume is the most commonly reported pathophysiologic impairment in post-acute effects of COVID-19, which is directly related to the severity of acute illness [31,32]. This observation is consistent with SARS and MERS and seems to be the contributing factor in long-term pulmonary sequelae of COVID-19. There is the viral-dependent invasion of endothelial-epithelial barrier causing infiltration of monocytes and macrophages, leading to extravasation of protein-rich exudate filling the alveolar space. This is similar to acute respiratory distress syndrome (ARDS) [33]. There are reports of pulmonary vascular micro and macrothrombosis in 20% of the patients with critical COVID-19 pneumonia and the severity of the endothelial injury and widespread microangiopathy seen on lung histopathology is greater than that seen in ARDS from other viruses [34,35].

Several other constitutional symptoms are demonstrated in this review [36-41]. The most important of them are weight loss, new-onset diabetes and hypertension, expectoration, blurred vision, and dry eyes. Chest pain is reported in up to 10% of COVID-19 survivors at 60 days follow up, while ongoing palpitations were reported in 8.53% at 6-months follow up. Apart from acute coronary syndrome (ACS) and myocarditis, an increased incidence of takotsubo cardiomyopathy is being reported in this pandemic compared with the pre-pandemic period (7.8% vs. 1.5%, respectively) [42]. Mechanisms contributing to cardiovascular sequelae in post-acute COVID-19 seem to be downregulation of ACE2 and renin-angiotensin-aldosterone system (RAAS), cytokine storm-related deterioration of myocardial integrity, pericarditis, and arrhythmias [43].

This systematic review had several limitations. One is the small number of studies with underpowered sample size, creating a potential bias and variation in defined outcomes leading to the heterogeneity of the results. Many studies used a self-reporting method which can produce an interobserver bias and almost all studies enrolled COVID-19 patients in mild, moderate, and severe disease category with variable follow-up times references. This can produce heterogeneous results. There was a predefined assessment of symptoms in every study assessed, which can lead to unreported outcomes. One other limitation is that there is no definition of the effect of one severity of COVID-19 and its associated symptoms. A critical illness survivor can have prolonged symptoms while a patient with mild disease can recover early from the same problem. Hence, there is a need for prospective studies to determine if the post-acute COVID-19 effects are a continuation of SARS-COV-2 or complications of premorbid conditions.

## Conclusion

The multiorgan sequelae of SARS-COV-2 infection beyond the acute infection are increasingly being recognized with an increasing clinical experience and pool of data becoming available rapidly on COVID-19. This updated systematic review of 21 studies and 54,730 patients is the largest cohort of patients with post-acute effects of COVID-19 evaluated to date. It demonstrated that post-acute effects of COVID-19 can persist even at six months and from the clinical point of view, medical professionals should look for the symptoms and signs in patients recovered from COVID-19. Necessary future research includes stratification of these post-acute effects with gender, age, and comorbid conditions in acute, subacute, and chronic phases of the disease. This will lead to a better understanding of the delayed sequelae of COVID-19. Through this review, it is clear that acute care of COVID-19 does not conclude at hospital discharge, and interdisciplinary care is needed for comprehensive care of these patients at homes and outpatient clinics. Hence, healthcare systems must establish dedicated COVID-19 clinics, where specialists from various disciplines can provide unanimous care.

## Data Availability

not applicable

