## Supplementary material for "Post-acute COVID-19 syndrome and its prolonged effects: An updated systematic review": supple 1

**NEWCASTLE - OTTAWA QUALITY ASSESSMENT SCALE**
**CASE CONTROL STUDIES**

Note: A study can be awarded a maximum of one star for each numbered item within the Selection and Exposure categories. A maximum of two stars can be given for Comparability.

### Selection

1) Is the case definition adequate?
   a) yes, with independent validation ✵
   b) yes, eg record linkage or based on self reports
   c) no description

2) Representativeness of the cases
   a) consecutive or obviously representative series of cases ✵
   b) potential for selection biases or not stated

3) Selection of Controls
   a) community controls ✵
   b) hospital controls
   c) no description

4) Definition of Controls
   a) no history of disease (endpoint) ✵
   b) no description of source

### Comparability

1) Comparability of cases and controls on the basis of the design or analysis
   a) study controls for _______________ (Select the most important factor.) ✵
   b) study controls for any additional factor ✵ (This criteria could be modified to indicate specific control for a second important factor.)

### Exposure

1) Ascertainment of exposure
   a) secure record (eg surgical records) ✵
   b) structured interview where blind to case/control status ✵
   c) interview not blinded to case/control status
   d) written self report or medical record only
   e) no description

2) Same method of ascertainment for cases and controls
   a) yes ✵
   b) no

3) Non-Response rate
   a) same rate for both groups ✵
   b) non respondents described
   c) rate different and no designation

**NEWCASTLE - OTTAWA QUALITY ASSESSMENT SCALE**
**COHORT STUDIES**

Note: A study can be awarded a maximum of one star for each numbered item within the Selection and Outcome categories. A maximum of two stars can be given for Comparability

### Selection

1) Representativeness of the exposed cohort
   a) truly representative of the average _______________ (describe) in the community ✵
   b) somewhat representative of the average ______________ in the community ✵
   c) selected group of users eg nurses, volunteers
   d) no description of the derivation of the cohort

2) Selection of the non exposed cohort
   a) drawn from the same community as the exposed cohort ✵
   b) drawn from a different source
   c) no description of the derivation of the non exposed cohort

3) Ascertainment of exposure
   a) secure record (eg surgical records) ✵
   b) structured interview ✵
   c) written self report
   d) no description

4) Demonstration that outcome of interest was not present at start of study
   a) yes ✵
   b) no

### Comparability

1) Comparability of cohorts on the basis of the design or analysis
   a) study controls for _____________ (select the most important factor) ✵
   b) study controls for any additional factor ✵ (This criteria could be modified to indicate specific control for a second important factor.)

### Outcome

1) Assessment of outcome
   a) independent blind assessment ✵
   b) record linkage ✵
   c) self report
   d) no description

2) Was follow-up long enough for outcomes to occur
   a) yes (select an adequate follow up period for outcome of interest) ✵
   b) no

3) Adequacy of follow up of cohorts
   a) complete follow up - all subjects accounted for ✵
   b) subjects lost to follow up unlikely to introduce bias - small number lost - > ____ % (select an adequate %) follow up, or description provided of those lost) ✵
   c) follow up rate < ____% (select an adequate %) and no description of those lost
   d) no statement
