## Supplementary material for "Post-acute COVID-19 syndrome and its prolonged effects: An updated systematic review": supple 2

### Appendix 2: Checklist for assessing quality of studies

Based on National Institutes of Health (NIH) study quality assessment tools for controlled trials (1).

| | **Yes** | **No** | **Other (CD, NR, NA)*** |
|---|---|---|---|
| 1. Was the study described as randomized, a randomized trial, a randomized clinical trial, or an RCT?<br>1.1. Was the method of randomization adequate (i.e., use of randomly generated assignment)? | | | |
| 2. Was the study described as a controlled trial?<br>2.2. Was the control group matched on relevant variables (age, gender, education, disorder)? | | | |
| 3. Was the overall drop-out rate from the study at endpoint 20% or lower of the number allocated to the intervention? | | | |
| 4. Was the differential dropout rate (between groups) at endpoint 15 percentage points or lower? | | | |
| 5. Was there high adherence to the intervention protocols for each treatment group? (defined as 75 % attendance or more) | | | |
| 6. Were other interventions avoided or similar in the groups? | | | |
| 7. Were outcomes assessed using valid and reliable measures? | | | |
| 8. Were outcomes measured consistently across all study participants? | | | |
| 9. Did the authors report that the sample size was sufficiently large to be able to detect a difference in the main outcome between groups with at least 80% power? | | | |
| 10. Were outcomes reported or subgroups analyzed prespecified (i.e., identified before analyses were conducted)? | | | |
| 11. For RCTs: were all randomized participants analyzed in the group to which they were originally assigned, i.e., did they use an intention-to-treat analysis?<br>11. For controlled studies: was a recognized statistical method employed? (recognized methods defined as dif-in-dif, regression discontinuity, propensity score matching, instrumental variables) | | | |
| **Total score: Number of YES** | | | |
| * CD: cannot determine; NR: Not reported; NA: Not applicable | | | |
| **Overall Quality Rating (Good, Fair, Poor)** | | | |
| Rater 1:<br>Rater 2: | | | |
| | Additional comments (If POOR, please state why): | | |

### References

(1) National Heart, Lung, and Blood Institute (NHLBI). Study Quality Assessment Tools. NHLBI. 2014. Available at: www.nhlbi.nih.gov/health-pro/ guidelines/in-develop/cardiovascular-risk-reduction/tools/
